# Co-Develop-IT - Co-design, Development, and Evaluation of Individually Tailored Technology-Enhanced Training and Rehabilitation Concepts: Methodological Guideline Development Study

**DOI:** 10.1101/2025.09.16.25335847

**Authors:** Patrick Manser, Lotte E.S. Hardeman, Andreas Wallin, David Moulaee Conradsson, Breiffni Leavy, Franziska Albrecht, Melvyn Roerdink, Jean-Jacques Temprado, Eling D. de Bruin, Erika Franzén

**Affiliations:** Division of Physiotherapy, Department of Neurobiology, Care Sciences, and Society, Karolinska Institutet, Huddinge, Sweden; Department of Human Movement Sciences, Faculty of Behavioural and Movement Sciences, Vrije Universiteit Amsterdam, Amsterdam Movement Sciences, Amsterdam, The Netherlands; Medical Unit Allied Health Professionals, Women’s Health and Allied Health Professionals Theme, Karolinska University Hospital, Stockholm, Sweden; Research and Development Unit, Stockholm Sjukhem Foundation, Stockholm, Sweden; NUTRIM Institute of Nutrition and Translational Research in Metabolism, Department of Nutrition and Movement Sciences, Faculty of Health, Medicine and Life Sciences, Maastricht University, 6211 LK Maastricht, The Netherlands; MHeNs Institute of Mental Health and Neurosciences, Department of Nutrition and Movement Sciences, Faculty of Health, Medicine and Life Sciences, Maastricht University, 6211 LK Maastricht, The Netherlands; Institut des Sciences du Mouvement (ISM), UMR 7287, CNRS, Aix Marseille Université, Marseille, France; Motor Control and Learning Group, Institute of Human Movement Sciences and Sport, Department of Health Sciences and Technology, ETH Zurich, Zurich, Switzerland; Department of Health, OST - Eastern Switzerland University of Applied Sciences, St. Gallen, Switzerland

**Keywords:** Community Participation, Digital Health, Disease Prevention, Exergaming, Health Promotion, Multidisciplinary Research, Participatory Research, Patient Participation, Rehabilitation, User-Centered Design

## Abstract

**INTRODUCTION:** Applying digital health technologies (DHTs) for health promotion and disease prevention is recommended by official bodies such as the World Health Organization. User-centered co-design with patient and public involvement is considered best practice for developing such complex interventions. While well-established frameworks are available to broadly guide such procedures, there is a need to better focus and guide the co-design process with preceding contextual research. Moreover, more specific guidance on additional methodological steps for the validation of DHT components and to facilitate their implementation, scalability, and sustainability would benefit the research community.

**OBJECTIVES:** This manuscript presents a consensus-based methodological guideline that delineates best practices for each step along the full lifecycle of DHT-enhanced training and rehabilitation concepts – from contextualization through co-development and evaluation to implementation.

**METHODS:** The *Co-Develop-IT guideline* was co-created through an expert consensus process. It consisted of biweekly 90-minute meetings between August 2024 and February 2025 in combination with written elaboration, feedback, and revisions between meetings, to gradually develop a consensus on best practice recommendations.

**RESULTS:** The *Co-Develop-IT guideline* consists of eight iterative phases directed toward multidisciplinary expert teams coordinating projects on DHT-enhanced training and rehabilitation concepts. It is applicable to any type(s) of end-user(s), exercise type(s), intended context(s) of use (e.g., primary healthcare, community health services, telemedicine) and overarching goal(s) (e.g., health promotion, primary through tertiary disease prevention). The guideline integrates and refines previous methodological frameworks and expands on existing best practices by introducing five distinct preparatory contextual research phases preceding generative co-design and development. These novel phases are dedicated to harmonizing all contributors’ interests and clarifying their level of involvement (particularly between research, industry, healthcare, and end-users), aligning co-design procedures with expectations and requirements of interest-holders, collaboratively delineating clear strategies to monitor project progression, and promoting implementation, scalability, and sustainability of the solutions to be co-developed.

**CONCLUSION:** The *Co-Develop-IT guideline* proposes best practices for how participatory research and patient and public involvement may be efficiently implemented and structured to benefit the establishment of individually tailored DHT-enhanced training concepts. Its main novelty lies in guiding the structured establishment of a more robust conceptual foundation through extensive preparatory contextual research phases, aimed at better targeting tailored co-development efforts toward successful implementation. We advocate for the continued refinement and consolidation of this methodological guideline to help the field strike a better balance between maximizing the benefits and mitigating the increased resource demands of collaborative research practices – ultimately maximizing its real-world impact.

## 1. Introduction

Innovations in digital health technologies (DHTs) – such as exergames or electronic and mobile health applications – have attracted significant interest as powerful tools to advance health promotion and disease prevention (primary through tertiary prevention initiatives; including rehabilitation) [1-4]. On the individual level, apart from promoting engagement and intervention adherence [2, 3], DHT-enhanced interventions can be designed to provide superior adherence to relevant principles of behavior-change, neuroscience, and exercise science compared to conventional exercise and rehabilitation modalities [2]. On a system’s level, DHTs stand out as scalable and cost-effective tools that can contribute to health system improvements via the implementation of such health promotion and disease prevention interventions in a well-controlled environment and hybrid intervention delivery [2, 4-6].

User-centered design and development methodologies, which continuously involve end-users and other multidisciplinary interest-holders in a participatory process, are considered best practice for designing and developing complex interventions such as DHT-enhanced interventions. Several frameworks have been published in recent years to guide these procedures. While a large number of studies failed to use guidelines to support rigorous intervention development, the UK Medical Research Council’s (MRC) guidance for developing and evaluating complex interventions [7] has been the most frequently used framework for developing rehabilitation interventions for older adults [8]. In the field of electronic health system development, many studies have referred to the use of participatory design methods without reference to a specific framework [9]. For the studies that have adopted a specific framework, User-Centered Design [10] has been the most frequently used one, albeit it has been adopted heterogeneously [9]. More specific guidance for healthcare contexts has recently been proposed with the generative codesign framework for healthcare innovation [11]. For DHTs specifically, the non-adoption, abandonment, scale-up, spread, and sustainability (NASSS) framework [12], the Multidisciplinary Iterative Design of Exergames (MIDE) - Framework [13], or its refined methodology from the ‘Brain-IT’ project [14] provide guidance on how to structure the conceptualization, co-design, and evaluation process.

The MIDE-framework [13] and its refined ‘Brain-IT’ methodology [14] have recognized that generative co-design can be better guided by setting clear boundaries on the freedom of this process to fulfill a project’s greater purpose. These boundaries are suggested to be based on a detailed elaboration of design requirements [13, 14] from the integration of findings from three pillars [14], which represented a paradigm shift in the approach to develop tailored DHT-enhanced interventions. These pillars include (1) existing empirical scientific evidence, together with research on the perspectives of (2) the intended primary end-users, as well as (3) the intended secondary end-users (e.g., healthcare professionals) and other relevant interest-holders (e.g., industry representatives) [14, 15].

While these methodological frameworks represented important steps to advance research methodologies, both were proposed by specific research groups, thus contrasting recommendations that such best practice recommendations or guidelines should be based on an expert consensus process [8]. Moreover, the application of the aforementioned methodologies to different projects (e.g., [15, 16]) has revealed room for further methodological advancements, which aligns with the observation that generic frameworks (such as MRC) were expanded to better fit project-specific contexts [8]. Specifically, there is a need for:

1. clearer recommendations to harmonize the interests and roles of different contributors – particularly across research, industry, healthcare, and end-users;
2. clarifying the level of involvement of different interest-holders;
3. additional methodological steps for the validation of DHT components and to facilitate the implementation, scalability, and sustainability of the solutions to be developed; and
4. harmonizing the interpretation and implementation of the recommended methodological steps and establishing a common terminology between different contributors, who often refer to similar issues with different wordings and theoretical references.

To address this need for further advancements of research methodologies, the research community would benefit from more rigorous methodological guidelines established based on an expert consensus process. These guidelines should delineate more specific, yet flexible enough, best practices for each step of contextualizing, co-designing, developing, evaluating, and implementing innovative rehabilitation interventions in general [8], and individually-tailored DHT-enhanced interventions specifically. To guide flexible and effective implementation in different projects [8], these guidelines would benefit from providing a checklist and application examples.

## 2. Objectives

The aim of this project was to co-create and present a consensus-based methodological guideline that delineates comprehensive best-practice recommendations for each step along the full lifecycle of individually tailored DHT-enhanced training concepts – from contextualization through co-development and evaluation to implementation.

To specifically address the need for consistent interpretation and application of this novel guideline, we additionally aimed to develop a corresponding checklist with item-specific explanations and elaborations to guide effective implementation. Finally, we aimed to briefly outline two application examples to demonstrate that the resulting guideline fulfills its primary purpose of providing more rigorous and specific, yet flexible enough best practice recommendations, and critically reflect on its relevance, novelty, and implications.

## 3. Methods

The project of co-creating this methodological guideline was initiated by the first author in June 2024. The *Co-Develop-IT guideline* was developed through an expert consensus process. It consisted of biweekly 90-minute meetings between August 2024 and February 2025 in combination with written elaboration, feedback, and revisions between meetings, to gradually develop a consensus on best practice recommendations. The application examples were worked out in the context of the projects ‘Park-MOVE’ and ‘Better Together’, which are being carried out at Karolinska Institutet, Sweden, and Vrije Universiteit Amsterdam, The Netherlands, respectively.

To provide a comprehensive methodological guideline that appropriately builds on previous research, the authors first exchanged in-depth information about their experiences in adopting existing guidelines and frameworks in their projects. We started with a critical discussion on which guidelines or frameworks were deemed useful to advance project methodologies and identified key areas for improvement (as summarized at the end of the Introduction). To develop more comprehensive and specific guidance, subsequent discussions and the consensus-finding process focused on which elements of well-established existing guidelines and frameworks can be integrated, revised, and, most importantly, be expanded to address the identified key areas for improvement.

## 4. Results

### 4.1 Overview

The *Co-Develop-IT guideline package* consists of the following complementary elements:

1. *Co-Develop-IT Core Guideline* (section 4.2)
2. *Explanation and elaboration sections* of the *Co-Develop-IT Core Guideline* (section 4.3)
3. *Co-Develop-IT Checklist* (Supplementary File 1)
4. Explanations and elaborations of each checklist item with guiding examples (Supplementary File 2)

#### 4.1.1 For which contexts are the *Co-Develop-IT guideline* intended?

The *Co-Develop-IT guideline* was developed for the context of individually tailored DHT-enhanced training and rehabilitation concepts that are purposefully co-developed with adequate theoretical underpinnings.

With *“DHT-enhanced training or rehabilitation concepts”*, we refer to:

i. a conceptual guideline that delineates all algorithmic decision trees regarding the structure, content, tailoring, delivery, supervision, and monitoring of a DHT-enhanced exercise, training, or rehabilitation program (in adherence with the definitions from [2, 17]), combined with:
ii. a purpose-developed DHT to implement and deliver the exercises, training, or rehabilitation program according to this concept.

For the sake of terminological simplicity, we will refer to *“DHT-enhanced training concepts”* from here on out.

With *“individually tailored”* training, we refer to integrating both *“personalization”* and *“individualized progression”*, understood as follows in the context of this guideline:

- *Personalization* refers to the systematic selection of the type, content, components, and characteristics of a training or rehabilitation program for a specific person based on their individual characteristics at baseline.
- *Individualized progression* refers to the gradual and systematic adaptation of training dose and stimuli to maintain a training load deemed sufficient to induce targeted health- or disease-related adaptations.

#### 4.1.2 Who is the *Co-Develop-IT guideline* intended for?

The *Co-Develop-IT guideline* is directed toward multidisciplinary expert teams coordinating projects involving the contextualization, co-design, development, evaluation, and implementation of individually tailored DHT-enhanced training concepts. It is applicable for any type(s) of end-user(s), exercise type(s), intended context(s) of use (e.g., primary healthcare, community health services, digital health), and overarching goal(s) (e.g., health promotion and disease prevention (primary through tertiary prevention; including rehabilitation)).

#### 4.1.3 How is the *Co-Develop-IT guideline* intended to be used?

To facilitate and support effective implementation of the conceptually discussed *Co-Develop-IT core guideline*, we provide the *Co-Develop-IT checklist* that details the exact steps to be performed and documented in projects following this guideline. We recommend that the checklist along with its item-specific explanation and elaboration sections be considered the core instruments to work with when implementing the guideline.

#### 4.1.4 What are key elements of the *Co-Develop-IT checklist*?

Apart from step-by-step instructions for the procedures to be completed and documented when adhering to the

*Co-Develop-IT checklist*, it defines the following *“contextual requirements*” for each item:

i. The minimum level of interest-holder involvement according to the participation choice points defined by Vaughn and Farrah (2020) [18] (contextually refined definitions for each level see heading of the *Co-Develop-IT checklist*).
ii. Which key interest-holders should be responsible for each item.
iii. Item relevance when adhering and reporting a project according to the *Co-Develop-IT guideline*.

#### 4.1.5 How does the *Co-Develop-IT guideline* balance rigor with practical flexibility?

The guideline’s development was centered around finding a good balance between methodological rigor and practical feasibility. Therefore, *“mandatory”* and *“recommended”* items are provided in the *Co-Develop-IT checklist*. While every project would benefit from adhering to all items, we acknowledge that, depending on the resources available, projects might not be able to adhere to all items. Therefore, we offer flexibility by the opportunity to choose a nuanced approach between the *“recommended”* best practice approach and a short-track option that only adheres to the *“mandatory”* items. The guidelines’ practical applicability, relevance, and flexibility are demonstrated by *two application examples* of the guideline in ongoing projects (Supplementary Files 3 and 4).

### 4.2 The Co-Develop-IT Guideline

The *Co-Develop-IT guideline* is structured in eight phases. An overview of its key structures and iterative steps that systematically guide projects towards making a real-world impact are illustrated in Figure 1. Importantly, both the guideline and each of the phases represent iterative project steps, meaning that a project moves up and down the phases – which may each involve multiple iterative loops – depending on goal achievement and project progress. A core component of the guideline is the establishment of project checkpoints with a traffic light system-based assessment framework with clear benchmarks and progression criteria. Thereby, a systematic and transparent approach to assess goal achievement is established, leading to iteratively looping a phase (= orange light) until the criteria to progress to the next phase (= green light) or regress to a previous phase to establish a more robust foundation (= red light) are met. Of course, this is also applicable to subprojects. t –

**Figure 1:**
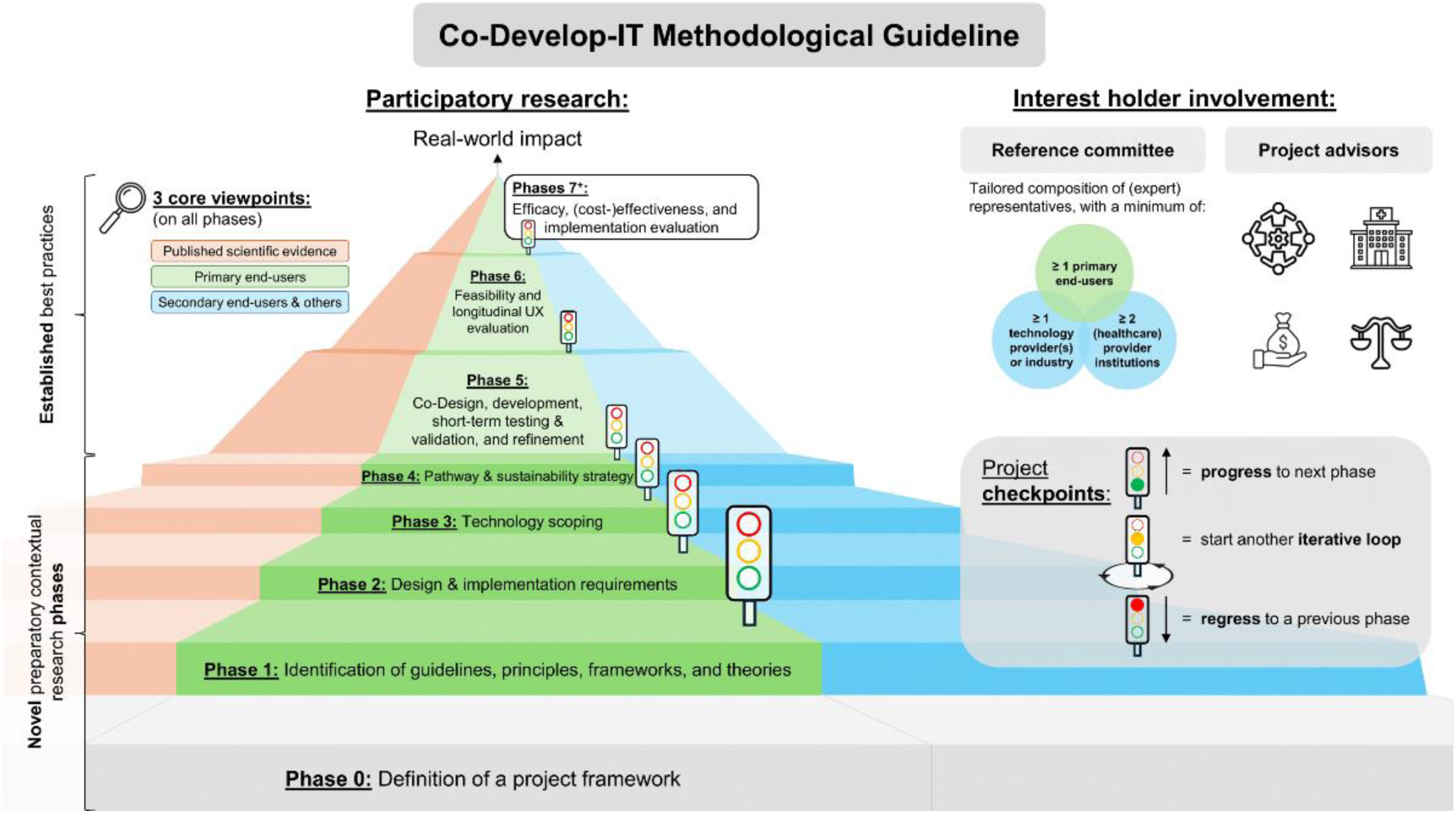
The *Co-Develop-IT guideline*. This figure illustrates an overview of the key components and phases of the *Co-Develop-IT guideline*, which systematically guides multidisciplinary expert teams coordinating projects on individually tailored technology-enhanced training concepts and related digital health technologies towards making a real-world impact. It is applicable to any type(s) of end-user(s), exercise type(s), intended context(s) of use (e.g., primary healthcare, community health services, telemedicine), and overarching goal(s) (e.g., health promotion, primary through tertiary disease prevention; including rehabilitation). Abbreviations: UX, user experience.

We acknowledge that details of specific steps and their order within a phase might not be applicable to every single project and might run to a certain extent in parallel. If necessary, details of specific steps or their order may be adapted, and these changes reported transparently. However, the order of completing each phase must be followed, as each phase builds on the findings of the previous phases.

Of note, all findings derived from projects adhering to the *Co-Develop-IT guideline* are to be reported following the appropriate checklist(s) provided on the EQUATOR (Enhancing the QUAlity and Transparency Of health Research) Network [19] to ensure reproducibility, transparency, and comparability to other research.

### 4.3 Explanation and Elaboration of the *Co-Develop-IT Guideline*

#### 4.3.1 Phase 0: Definition of Project Framework

The initial phase of the process of applying the *Co-Develop-IT guideline* is dedicated to:

i. defining *non-negotiable key elements* of the project (i.e., Phase 0.1);
ii. defining *requirements on interest-holder involvement*, resources, and the regulatory environment for the project (i.e., Phase 0.2);
iii. *establishing a project consortium* including all relevant interest-holders and *defining their roles and responsibilities* throughout the project for effective collaboration and accountability (i.e., Phase 0.3); and,
iv. collaboratively agreeing on specific *project checkpoints* and a preliminary *project time plan* (i.e., Phase 0.4).

These steps are to establish a collaboratively agreed-upon framework within which the project should take place.

##### Phase 0.1: Define Overall Context and Goals of the Project

Initiate the project by outlining its:

✓ *overarching goals and broader intended context* (*Co-Develop-IT checklist* ☑-item 3; e.g., which structures or functions the training should impact, type and multidisciplinary expertise of involved institutions),
✓ *target population(s)* (☑-item 4; e.g., individuals at risk for or living with a specific disease or condition as primary end-users along with healthcare professionals and relatives/supporters as secondary end-users,
✓ *intervention type* (☑-item 5; e.g., DHT-enhanced multidomain (physical, motor, and cognitive) training), and
✓ *targeted outcome domain* (☑-item 6; e.g., health-related quality of life).

Outlining these aspects allows delineating a clear path to follow, sets clear boundaries for the freedom of the subsequent co-development process toward fulfilling a project’s larger purpose, and guides the identification of relevant interest-holders to be recruited (phase 0.2). Therefore, make sure these definitions are backed on a clearly defined theoretical rationale for how the development of an individually tailored DHT-enhanced training concept can address a specific problem or knowledge gap (☑-items 1 and 2). Leave sufficient room for innovations and novel ideas that may arise during the co-development process. To do so, no specifics such as exercise or training variables, specific DHTs to be used to implement the training, or any design elements of the DHTs and the training are to be defined at this stage. Once a project consortium is established (phase 0.4), these initial definitions are to be refined and agreed on collaboratively with all relevant interest-holders (☑-item 14) – thereby providing a structured approach to ensure that the overall project context and goals are backed both by both a theoretical rationale and real-world patient and public needs.

##### Phase 0.2: Define Requirements on Interest-Holder Involvement, Resources, and Regulatory Environment for the Project

In the second subphase, identify:

i. contextual requirements and required multidisciplinary interest-holders (☑-item 7),
ii. required resources for each interest holder (☑-item 8; e.g., time, staff, funding, expertise, facilities, and data resources), and
iii. regulatory requirements (☑-item 9) to successfully complete the project (e.g., institutional, ethics, and good clinical practice regulations, and requirements for collaboration agreements).

Table 1 provides a list of potential interest-holders to be considered to ensure that the project is grounded in a realistic and competent environment.

**Table 1:**
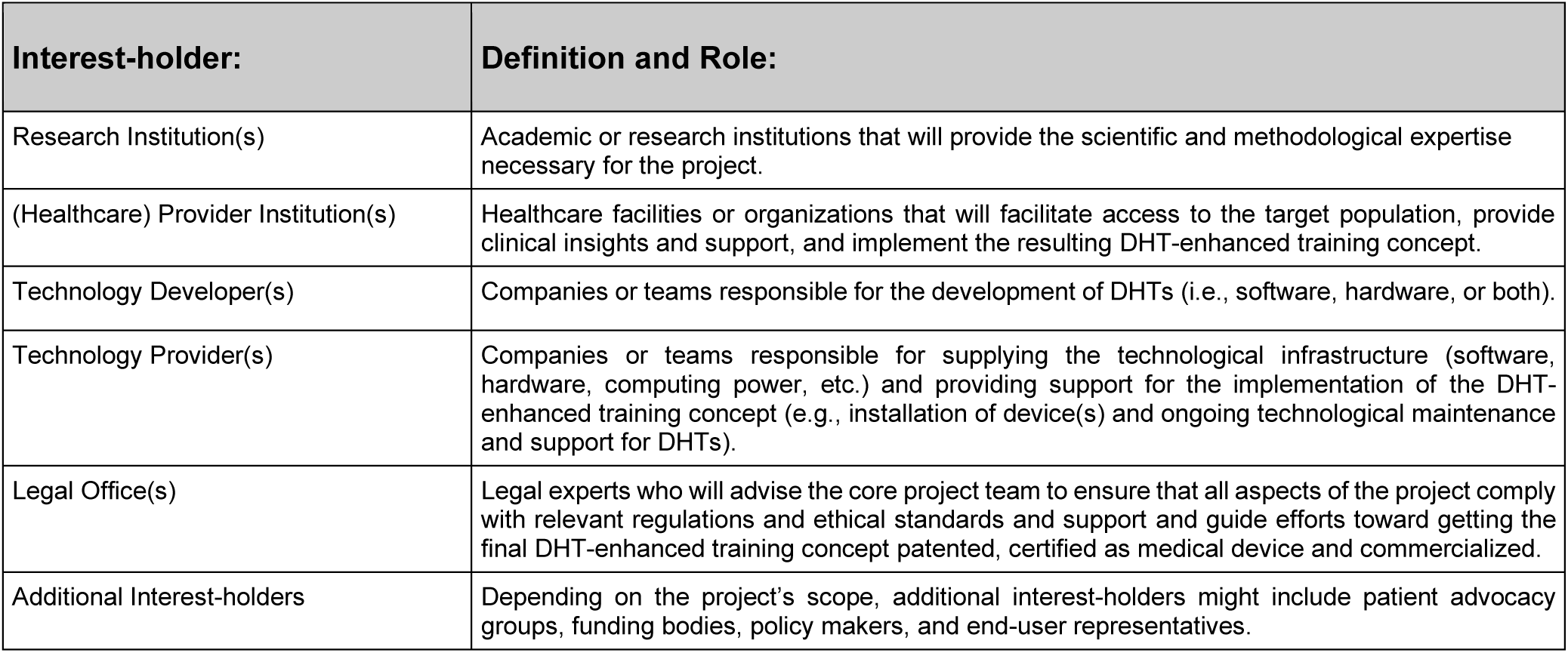
Definitions and key role of multidisciplinary interest-holders to be considered to be involved in a research projects in the co-design, development, and evaluation of individually tailored DHT-enhanced training concepts. Abbreviations: DHT, digital health technology;

| Interest-holder: | Definition and Role: |
| --- | --- |
| Research Institution(s) | Academic or research institutions that will provide the scientific and methodological expertise necessary for the project. |
| (Healthcare) Provider Institution(s) | Healthcare facilities or organizations that will facilitate access to the target population, provide clinical insights and support, and implement the resulting DHT-enhanced training concept. |
| Technology Developer(s) | Companies or teams responsible for the development of DHTs (i.e., software, hardware, or both). |
| Technology Provider(s) | Companies or teams responsible for supplying the technological infrastructure (software, hardware, computing power, etc.) and providing support for the implementation of the DHT-enhanced training concept (e.g., installation of device(s) and ongoing technological maintenance and support for DHTs). |
| Legal Office(s) | Legal experts who will advise the core project team to ensure that all aspects of the project comply with relevant regulations and ethical standards and support and guide efforts toward getting the final DHT-enhanced training concept patented, certified as medical device and commercialized. |
| Additional Interest-holders | Depending on the project's scope, additional interest-holders might include patient advocacy groups, funding bodies, policy makers, and end-user representatives. |

Moreover, include legal experts at this preparatory stage to outline the steps required to patent the final training concept or obtain medical device certification for the DHTs (i.e., software, hardware). This will ensure that all relevant aspects are considered during the project. This may include adhering to standards set by regulatory bodies such as the Food and Drug Administration in the United States or the European Medicines Agency in Europe. The recently published Common European Classification Grid for digital medical devices is helpful to facilitate the definition of the taxonomy and evidence requirements for assessment of DHTs and offers the possibility to have a common reference on a European level against which national classifications and evidence requirements can be mapped [20]. Key considerations include device classification, safety and efficacy testing, cost-effectiveness evaluation, data security, labeling, and post market surveillance.

##### Phase 0.3: Establish a Project Consortium and Define each Interest-holders’ Roles Throughout the Project

In the third subphase, recruit interest-holders to establish a project consortium based on the drafted project goals and context from subphase 0.1 and the requirements for a complete project consortium defined in phase 0.2. Ensure that at least one interest-holder representing each target population (☑-item 4) and multidisciplinary context (☑-item 7) is involved.

Establishing a project consortium entails setting up a *project reference committee* (☑-item 10b; mandatory item) and including *project advisors* (☑-item 10c; recommended item) to continuously guide the *core project team* (☑- item 10a; mandatory item). In the case of multinational projects, establish a project reference team and recruit project advisors for each country involved to ensure that local and socio-cultural nuances are sufficiently considered. Tailor the project consortium’s constellation to the specific research project, adhering to the minimum key interest-holders and responsibilities listed in Table 2. Of note, the involvement of members of the project consortia might change throughout the conduct of a project, depending on the project-specific requirements, needs, or relevance in different phases.

**Table 2:**
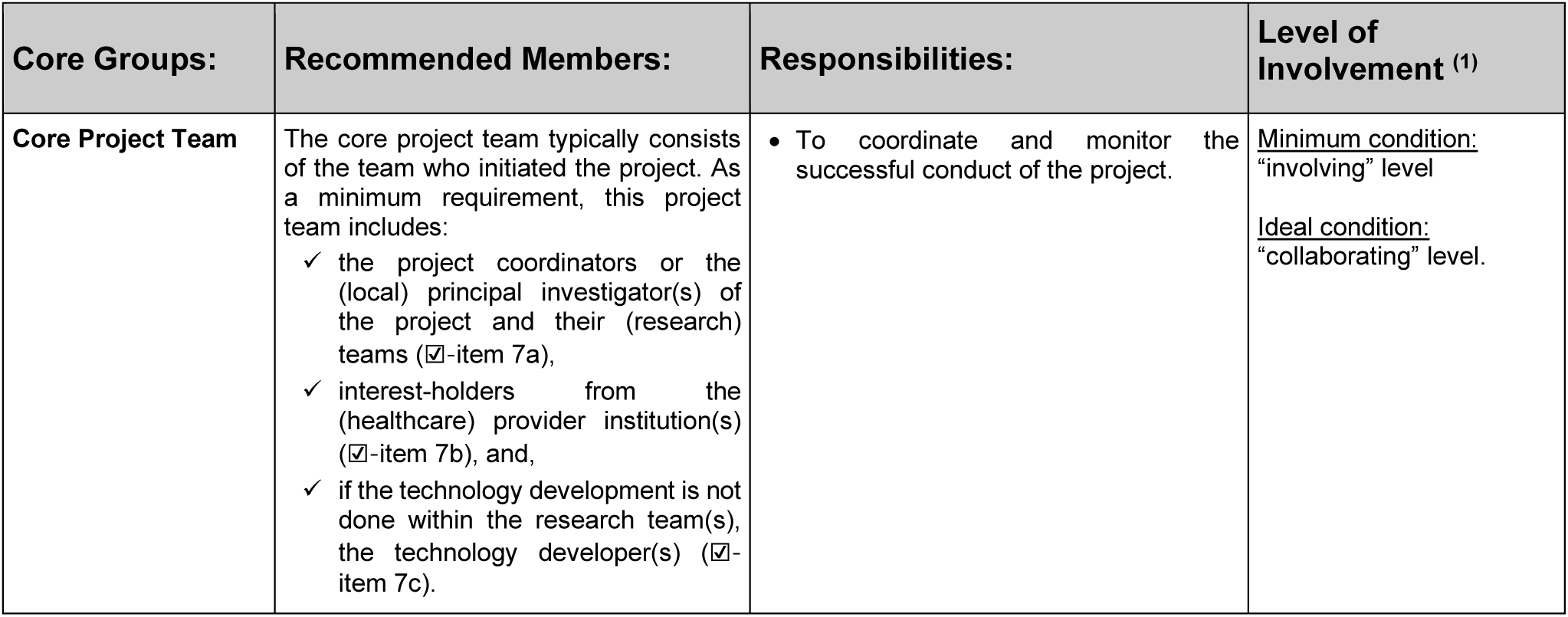
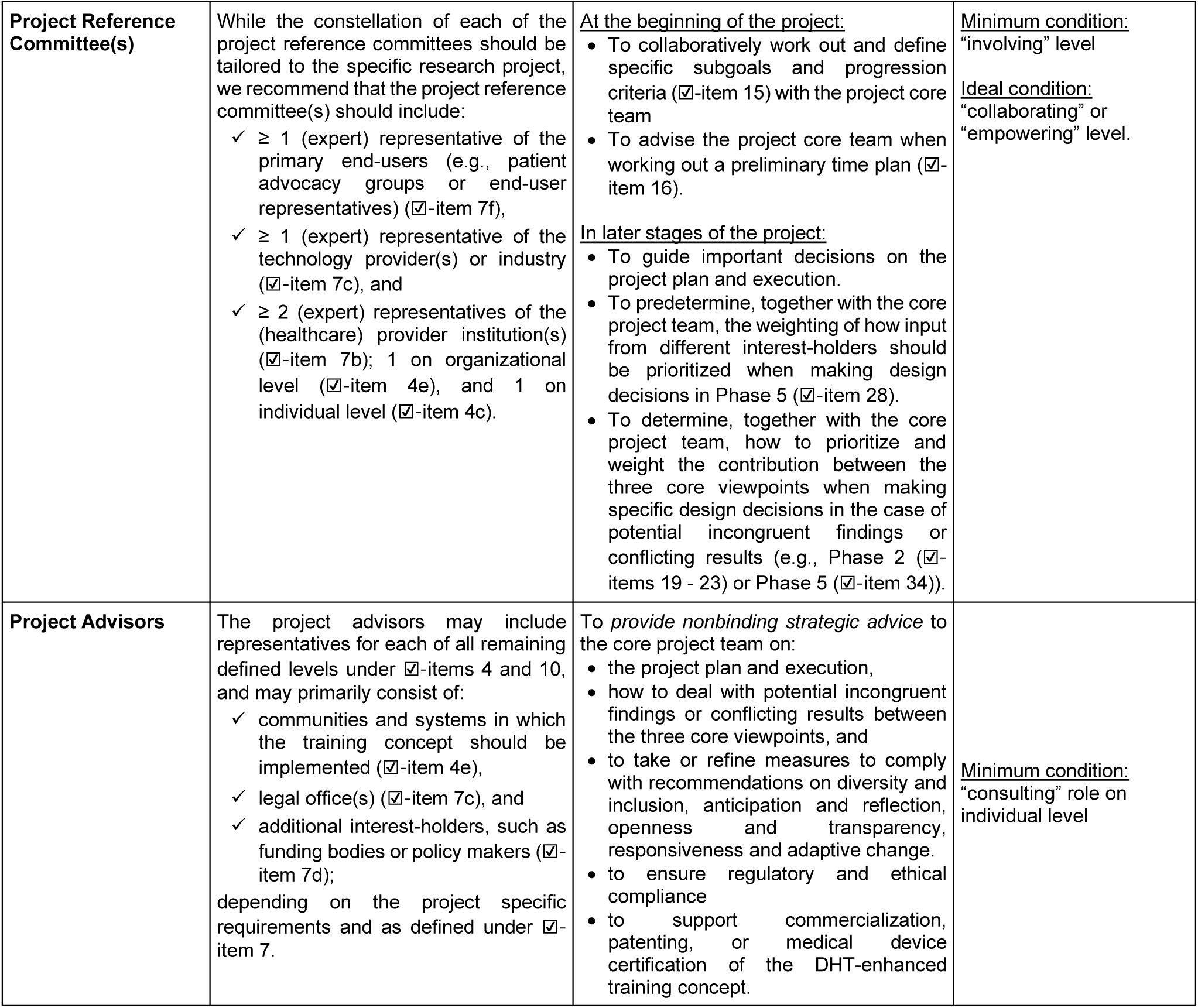
Recommendations on key interest-holders and responsibilities of core groups to be involved in research projects following the *Co-Develop-IT guideline*. (1) Level of participatory research involvement according to the participation choice points outlined by Vaughn and Farrah (2020) [18]. Abbreviations: DHT, digital health technology.

| Core Groups: | Recommended Members: | Responsibilities: | Level of Involvement <sup>(1)</sup> |
| --- | --- | --- | --- |
| **Core Project Team** | The core project team typically consists of the team who initiated the project. As a minimum requirement, this project team includes:<br>✓ the project coordinators or the (local) principal investigator(s) of the project and their (research) teams (☑-item 7a),<br>✓ interest-holders from the (healthcare) provider institution(s) (☑-item 7b), and,<br>✓ if the technology development is not done within the research team(s), the technology developer(s) (☑-item 7c). | • To coordinate and monitor the successful conduct of the project. | <u>Minimum condition:</u><br>"involving" level<br><u>Ideal condition:</u><br>"collaborating" level. |
| **Project Reference Committee(s)** | While the constellation of each of the project reference committees should be tailored to the specific research project, we recommend that the project reference committee(s) should include:<br>✓ ≥ 1 (expert) representative of the primary end-users (e.g., patient advocacy groups or end-user representatives) (☑-item 7f),<br>✓ ≥ 1 (expert) representative of the technology provider(s) or industry (☑-item 7c), and<br>✓ ≥ 2 (expert) representatives of the (healthcare) provider institution(s) (☑-item 7b); 1 on organizational level (☑-item 4e), and 1 on individual level (☑-item 4c). | <u>At the beginning of the project:</u><br>• To collaboratively work out and define specific subgoals and progression criteria (☑-item 15) with the project core team<br>• To advise the project core team when working out a preliminary time plan (☑-item 16).<br><u>In later stages of the project:</u><br>• To guide important decisions on the project plan and execution.<br>• To predetermine, together with the core project team, the weighting of how input from different interest-holders should be prioritized when making design decisions in Phase 5 (☑-item 28).<br>• To determine, together with the core project team, how to prioritize and weight the contribution between the three core viewpoints when making specific design decisions in the case of potential incongruent findings or conflicting results (e.g., Phase 2 (☑-items 19 - 23) or Phase 5 (☑-item 34)). | <u>Minimum condition:</u><br>"involving" level<br><u>Ideal condition:</u><br>"collaborating" or "empowering" level. |
| **Project Advisors** | The project advisors may include representatives for each of all remaining defined levels under ☑-items 4 and 10, and may primarily consist of:<br>✓ communities and systems in which the training concept should be implemented (☑-item 4e),<br>✓ legal office(s) (☑-item 7c), and<br>✓ additional interest-holders, such as funding bodies or policy makers (☑-item 7d);<br>depending on the project specific requirements and as defined under ☑-item 7. | To *provide nonbinding strategic advice* to the core project team on:<br>• the project plan and execution,<br>• how to deal with potential incongruent findings or conflicting results between the three core viewpoints, and<br>• to take or refine measures to comply with recommendations on diversity and inclusion, anticipation and reflection, openness and transparency, responsiveness and adaptive change.<br>• to ensure regulatory and ethical compliance<br>• to support commercialization, patenting, or medical device certification of the DHT-enhanced training concept. | <u>Minimum condition:</u><br>"consulting" role on individual level |

To ensure project feasibility and mitigate potential risks of non-completion, comprehensive resource mapping and comparing the available resources for all interest-holders (☑-item 11) with the required resources (☑-item 8) helps to identify any shortcomings to be addressed to support smooth project execution and completion (☑-item 12). Critically reflect on any relevant additional project-specific resources and risks to ensure thorough risk mitigation.

Establish formal collaboration agreements that delineate the responsibilities, rights, obligations, and degree of involvement of each collaborator (☑-item 13) to facilitate the management of expectations and ensure accountability. Regularly revisit and update the collaboration agreements as needed to reflect any changes in the project scope or interest-holder roles.

##### Phase 0.4: Collaboratively Agree on Specific Subgoals and Progression Criteria as well as a Preliminary Time Plan for the Project

Finally, collaboratively refine and agree on the specific goals and subgoals of the project (☑-items 14 and 15a; as drafted in phase 0.1) and define *project checkpoints* (☑-item 15) that are aligned with the overall goals of the project and its phases – both based on a consensus process with all interest-holders of the project consortium (i.e., core project team and reference committee and (optionally) project advisors). Each project checkpoint requires a traffic light system-based assessment framework with clear benchmarks (☑-item 15c) and progression criteria (☑-item 15d) to ensure transparency in the assessment of agreed-upon goals (☑-item 15b). The project checkpoints are usually to be positioned towards the end of each phase inform whether the project:

✓ can progress to the next phase (= green light),
✓ requires further iterative loops with refinements before proceeding to the next phase (= orange light), or
✓ requires the establishment of a more robust foundation by regressing to a previous phase (= red light).

Descriptive examples are provided in Supplementary File 2 or in previous literature of the author team in relation to feasibility and user experience testing in preparation for a randomized controlled trial (RCT) [21, 22]. For transparency in the interpretation of project results and progression, we recommended that the traffic light system-based assessment framework be published (as part of a project protocol) or at least pre-registered (e.g., on the Open Science Framework) prior to the start of data collection.

Collaboratively agreeing on these goals and checkpoints establishes a unified direction and commitment to the project’s success and helps prevent the emergence of misunderstandings or conflicts at a later stage of the project. To support these efforts, adopt techniques such as empathetic target group analysis [23] to foster a better understanding of the diverse needs and perspectives among interest-holders before starting the co-design or co-creation process [24, 25]. Moreover, have the added value and expectations of each partner in collaboratively achieving these goals communicated to transparently foster fruitful collaboration.

Finally, collaborative development and an agreement on a *preliminary project time* (☑-item 16) plan are essential to align interest-holder expectations regarding progress and output per unit of time. This is particularly important for harmonizing the expected rates of progress in public-private partnerships, given that public research institutions may be perceived by the industry as slow-moving ‘oil tankers’, whereas the industry may be seen by academia as agile ‘speedboats’ rushing toward marketable outputs.

#### 4.3.2 Phase 1: Identification of Guidelines, Principles, Frameworks, and Theories

Phase 1 is dedicated to identifying guidelines and evidence-based recommendations for the overall goals of the project (☑-item 17) along with principles, frameworks, or theories (☑-item 18). This phase is key to defining a robust backbone and guiding future steps of projects, particularly decisions on the design, characteristics, and content of the DHT-enhanced training concept to be developed. A few broadly applicable examples include:

##### Guidelines and evidence-based recommendations

✓ clinical or best practice guidelines for a specific target population
✓ consensus statements, such as the global consensus for optimal exercise recommendations for enhancing healthy longevity in older adults [26]

##### Principles

✓ general training principles [17, 27]
✓ neuroplasticity principles [28, 29]
✓ principles for neurorehabilitation (i.e., that integrate motor learning and brain plasticity mechanisms) [30]

##### Frameworks

✓ a behavior-change framework applicable to physical activity or training [31]
✓ guided plasticity facilitation framework [32] for motor-cognitive training
✓ the adaptive capacity model [33]
✓ the *‘Beyond “Just” Fun of Exergames - Framework’* [2]
✓ implementation and upscaling frameworks like NASSS [12] or PRACTIS (the PRACTical planning for Implementation and Scale-up) [34].

##### Theories

✓ Theory of Effort Minimization in Physical Activity [35, 36]
✓ OPTIMAL (Optimizing performance through intrinsic motivation and attention for learning) theory of motor learning [37]
✓ A behavior-change theory supporting initiation and consolidation of physical activity behavior changes [38]

#### 4.3.3 Phase 2: Determine Design and Implementation Requirements

The overarching aim of Phase 2 is to delineate a comprehensive set of design and implementation requirements for the DHT-enhanced training concept. This encompasses a range of considerations to be adhered to throughout the subsequent co-development phases, including:

i. generating *user models* (☑-item 19);
ii. defining requirements for *core components* of individually tailored training concepts (☑-item 20);
iii. defining *environmental requirements* (e.g., training equipment, space requirements, connectivity such as Wi-Fi or Bluetooth; ☑-item 21);
iv. defining *hardware* (☑-item 22) *and software* (☑-item 23) *requirements* for the DHTs.

Elaboration of the design and implementation requirements should be built on Phase 1 by integrating identified guidelines and evidence-based recommendations (☑-item 17) and taking into account relevant principles, frameworks, and theories (☑-item 18). Consider the set of core question provided in Supplementary File 5 and the checklist facilitating DHT adoption provided by Hamasaki et al. (2025) [39] to ensure that all potentially relevant aspects are considered. All decisions are to be made by integrating the findings from three core viewpoints:

1. A synthesis of published *scientific evidence*; together with the findings from performing qualitative research on:
2. the perspectives of the *intended primary end-users*, and
3. the *intended secondary end-users* and other relevant interest-holders.

The *current state of the scientific evidence* should be derived from conducting an umbrella review, meta-analysis, systematic, scoping, or narrative review based on the established levels of evidence [40]. If these types of high-level evidence were already reported in recent scientific literature, a narrative synthesis of the current state of evidence suffices. The *perspectives from intended primary and secondary end-users and all other relevant interest-holders* should be derived from conducting qualitative research (based on, e.g., semi-structured interviews or focus groups). Refer to ☑-item 29 on how to set the stage to facilitate reflective dialog and productive workshops and apply these recommendations also to the qualitative research components in this phase (☑-items 19 - 23).

We recommend conducting individual interviews with primary end-users separately from the focus group discussions with the remaining interest-holders to align data collection to reflect the three core viewpoints from which insights to derive decisions on the design and implementation requirements are to be combined. To provide a more nuanced understanding of how inclusivity and accessibility can be ensured for all relevant intended end-users, consider examining the current state of evidence and interview/focus group transcripts through the lens of critical discourse analysis [41]. These insights should be aligned with broader frameworks for sustainable innovation and ethical design in digital health [42].

#### 4.3.4 Phase 3: Technology Scoping

The third phase focuses on identifying (☑-item 24a) and critically appraising (☑-item 24b) existing (partial) DHT solutions in both research and the market that align with the project’s overall goals (Phase 0). Apply techniques such as trend analysis and the SWOT (acronym for Strength, Weakness, Opportunities, and Threats) matrix [13] to gain a comprehensive understanding of the current state of knowledge and to identify areas for further improvement. Moreover, assess each identified solution against the design and implementation requirements defined in Phase 2 (☑-item 24c). Understanding the strengths, limitations, and areas for improvements of current solutions in relation to the design and implementation requirements is essential for building robust foundation for advancing the field with credible, innovative, and evidence-based DHT-enhanced training concepts. Finally, identify and analyze emerging trends (☑-item 24d) to ensure that the project identifies and integrates relevant technological advancement that may be relevant to better fulfill the design and implementation requirements. This is especially important for rapidly evolving technological opportunities, such as augmented or mixed reality, artificial intelligence, bio- or neurofeedback, and brain-computer interfaces [2].

#### 4.3.5 Phase 4: Define Pathway & Sustainability Strategy

The aim of Phase 4 is to make a strategic decision on the project’s development path (☑-item 25) and establish a sustainability strategy for the DHT-enhanced training concepts to be (further) developed (☑-item 26). Specifically, on the basis of the findings of Phases 1 - 3, determine whether the project strives to develop a novel DHT-enhanced training concept from scratch (*Path 1*) or further develops and builds on existing solutions (*Path 2*).

A purpose-developed software builds the heart of every DHT, whereas the conceptual decisions and algorithmic decision trees provided in a training concept build the heart of a DHT-enhanced training concept [2]. Therefore, to maximize scalability and transferability to other application scenarios or use cases, focus on developing training concepts and software to implement the training while relying on (i) well-established, off-the-shelf hardware and (ii) ensuring the software is universally applicable with different hardware peripherals. This adheres to the *“training [instead of product] first”* approach [43] and facilitates the sustainability strategy, as it reduces the complexity of providing necessary equipment and improves scalability.

In this regard, under ☑-item 26, a sustainability strategy is to be developed to ensure that the solution (DHT-enhanced training concept) to be (further) developed will be made available to the intended target populations and remain available after the project’s completion. This strategy should address its long-term availability, which could be provision and maintenance of open-access to software, developing a business plan and outlining a plan to commercialize the solution, or transferring intellectual property rights to an existing company. Depending on this, mechanisms for ongoing maintenance and support, scalability to reach a broader audience or adapt to different contexts, and identifying resources to support the long-term sustainability of solutions are to be worked out. This step is crucial to ensure that the project’s outcomes have a lasting impact and continue to benefit the target populations in the long-term.

With these extensive preparatory contextual research steps from Phases 1 - 4, a robust conceptual foundation for the targeted co-development towards successful implementation phases of the project is laid.

#### 4.3.6 Phase 5: Co-Design, Development, Short-Term Testing & Validation, and Refinement

##### Phase 5 consists of multiple iterative cycles of

i. *co-designing and developing* prototypes of the DHT-enhanced training concept (Phases 5.2 to 5.5),
ii. short-term user experience *and safety testing* and *validation* of components of the DHT-enhanced training concept (Phase 5.5), and
iii. *refining* these *prototypes* by further co-design and development (Phase 5.6).

This iterative cycle is to be repeated until an *“acceptable”* solution (= all components of the DHT-enhanced training concepts have been successfully validated, have a good user experience, and are safe) is achieved. The result of this phase is an *“original”* DHT-enhanced training concept that enters the next phases of longitudinal evaluations.

##### Phase 5.1: Build a Framework for the Co-Design

First, identify congruent aspects from previous project phases (☑-item 27a) and agree on which of the remaining aspects for the DHT-enhanced training concepts require co-design procedures (☑-item 27b). Subsequently, work out and define, together with the project reference committee(s), the weighting of how inputs from different interest-holders is to be prioritized when collaboratively making design decisions (☑-item 28). For example, it might be agreed that design decisions on the graphical user interface should primarily be driven by primary and secondary end-users, whereas algorithms for individualized tailoring of the training should mainly rely on scientific evidence with input from researchers and healthcare professionals. Clear definitions of each interest-holder’s role and expected contributions helps streamline and guide the complex task of integrating various, potentially conflicting perspectives and viewpoints from different interest-holders – increasing the likelihood of successfully generating conceptual prototypes that fulfill all the design and implementation requirements defined in Phase 2 of the project and all involved contributors can agree on.

##### Phase 5.2: Co-Design Workshops

The diversity in background, education levels, and experiences of interest-holders can be expected to lead to power dynamics that influence or limit the interactions during the co-design process [44]. Therefore, a stepwise process is recommended to set the stage for a well-functioning collaborative effort.

Specifically, start the co-design workshops with a short presentation to share: the rational and overall goals of the project (☑-item 29a); preparatory findings from earlier phases of the project (☑-item 29b); and clarify the planned procedures (☑-item 29c) and expectations (☑-item 29d) with all co-design workshop participants. With respect to the latter, transparently share the weighting and prioritization of input from different interest-holders (☑-item 28) so that all participants have a clear understanding of their expected contributions while mitigating potential power imbalances. Finally, invest time in establishing mutual trust, empathy and comfort with all participants (☑-item 29e). This step is critical to foster a better understanding of the diverse needs and perspectives among interest-holders and facilitate reflective dialogue before starting the co-design or co-creation process [24, 25]. Employ techniques such as empathetic target group analysis [23] to achieve this aim.

If the project chooses path 2 – to further develop existing DHT-enhanced training concepts (☑-item 27) – present these solutions to all focus group participants to provide them with a clear understanding of what the project builds on and what preparatory steps led to the chosen starting point (☑-item 30). The participants should also be given the opportunity to try these existing DHTs to help them identify opportunities for improvements that can be addressed in generative co-design workshops.

Finally, the *“generative”* part of co-design is performed (☑-item 31). During the co-design process, employ techniques such as 6-3-5 brainwriting [45] to ensure that all interest-holders can express their ideas without judgment [24, 25]. To facilitate collaborative idea generation, use different paper- and pencil-based, embodied, or technology-supported prototyping techniques. Simple methods may include, but not limited to, empathy mapping [23, 46], user journey mapping [47], bodystorming [48, 49], collaborative sketching and drawings for the rapid development of initial concepts and designs [13], or – when building on and further developing existing solutions – the cognitive walkthrough method [50]. Technology-enhanced prototyping may include the use of middleware interfacing software tools that allow for the rapid development of initial concepts and designs to the creation of prototypes of the proposed concepts (e.g., game mechanics, theme) in game engines. Such tools allow early identification of possible modifications that must be performed to facilitate user interaction (e.g., removing the need to press buttons) and are particularly useful for the development of gamified DHTs. [13]

##### Phase 5.3 and 5.4: Data Synthesis and Development

Analyze data generated during the *“generative”* part of co-design, such as audio (and video) recordings combined with generated sketches and drawings or virtual conceptual prototypes, using qualitative research methodologies (☑-item 32), such as qualitative content analysis [51-53]. Use the frameworks defined in phase 1 to guide the interpretation of findings. For example, the Co-Design Evaluation Framework could be useful in this stage to ensure appropriate consideration of the impacts of different contextual considerations [54].

In the next step, present the results from each workshop to the project reference committee to collaboratively integrate the findings and rank-order complementary solutions. It is likely that there are incongruent findings or conflicting results between different workshops and complementary solutions for the same aspects of the DHT-enhanced training concepts from different workshops. In this case, present these results to the project reference board(s) and collaboratively make consensus-guided decisions on how to integrate the findings into decisions on the design of the DHT-enhanced training concepts. Ideally, consider the advice of project advisors as well.

Based on the derived rank-ordering (☑-item 33) and considering resource availability (☑-item 11), derive a list of development tasks (☑-item 34) and develop testable prototypes of the DHT-enhanced training concept (☑-item 35).

##### Phase 5.5: Short-Term Testing and Validation

The first prototypic components of the DHT-enhanced training concepts are then thoroughly tested on safety and user experience (☑-item 36), and relevant components of the DHT-enhanced training concept are validated (☑-item 37).

User experience refers to a complex characteristic that results from the perception of many distinct quality aspects of a product [55]. The relevance of these quality aspect varies between different contexts and products under evaluation [56-58]. It has been defined as a person’s perceptions and responses resulting from the use and anticipated use of a product, system, or service [59]. For a comprehensive user experience assessment, combine the advantages of both quantitative and qualitative research methodologies. Consider Campbell’s (2024) [60] step-by-step instructions and best practices.

For *quantitative user experience evaluations*, research has often relied on questionnaires for specific subconstructs of user experience, such as usability, which is a prominently evaluated subconstruct in the field of DHTs [58]. For a more comprehensive user experience assessment, we recommend the validated modular framework introduced by Schrepp and Thomaschewski (2019) [55]. This framework contains several scales that measure different user experience aspects and allows the construction of customized questionnaires tailored to the specific research question and context of use in a uniform format. While the shortcoming of such a modular questionnaire is the lack of standardized benchmarks, its application is mainly recommended when multiple measurements of the same product are compared over time [55], matching the iterative process proposed by the *Co-Develop-IT guideline*. This scale may be complemented with other – more specific – scales for certain new use cases and product types to provide a more comprehensive assessment (examples see item-specific explanation and elaboration statement) [55]. For *qualitative user experience evaluations*, obtain data complementary to quantitative evaluations to inform further developments and refinements. Specifically, collect specific suggestions for improvements in the components of the DHT-enhanced training concepts to optimize the user experience via semi-structured interviews with primary and secondary end-users.

Assess the *safety* of components of the DHT-enhanced training concepts and interaction with DHTs in relation to the characteristics and capabilities of the targeted end-users. Consider the perspectives from both primary and secondary end-users. We recommend having trained professionals closely observing play-test sessions to assess safety (e.g., risk of falls or injuries) along with qualitative research with primary and secondary end-users to assess perceived safety and collecting suggestions for improvements.

Thoroughly *validate* the relevant components of the DHT-enhanced training concepts according to the requirements defined under ☑-item 15b. This may include but is not limited to software algorithms for player movement detection (e.g., [61]), (game) performance metrics (e.g., [62-66]), (gamified) assessments (e.g., [67]), or mechanisms for individualized tailoring (e.g., [62]).

At the end of this subphase, check whether the quality criteria defined under 15c are reached (☑-item 38), share the findings with all co-design workshop participants (☑-item 39), and progress according to the mechanism outlined under ☑-item 15d.

##### Phase 5.6: Iterative Refinements

Iteratively repeat subphases 5.1 to 5.5 until an *“acceptable”* solution is achieved (☑-item 40) according to the specific subgoals and quality criteria defined under ☑-items 15a and 15c. Once this has been achieved, progress with longitudinal evaluations.

#### 4.3.7 Phase 6: Feasibility and Longitudinal User Experience Evaluation

For the phases introduced thus far, the checklist provides best practice recommendations that are novel, expand existing guidelines or frameworks, and are specific to DHT-enhanced training. Subsequent phases 6 and 7+, covering longitudinal evaluations and implementation, are more generally applicable and well covered by established guidelines. Therefore, we provide (i) recommendations which established procedures to follow and (ii) which additional elements to consider to make these guidelines better tailored to DHT-enhanced training.

In phase 6, test the feasibility of the full DHT-enhanced training concept and the study procedures for subsequent full-scale trials, along with a more in-depth investigation of the user experience and safety of the full DHT-enhanced training concept (☑-item 41 and 42). This phase is again iterative, meaning that it is repeated according to prespecified progression criteria until a solution with acceptable feasibility and user experience among primary and secondary end-users is achieved (☑-item 43).

Evaluate *feasibility* of the study procedures and the DHT-enhanced training along with its *user experience*. We recommend to do these evaluations on the basis of the conceptual framework of Eldridge et al. (2016) [68], following the terminology and recommendations of the Medical Research Council guidance [7], and considering mixed-method evaluations of both feasibility and user experience following O’Cathain et al. (2015) [69] and Campbell (2024) [60]. Like in Phase 5, collect complementary qualitative data that provides broader contextual information and obtain data on specific suggestions for improving the feasibility and user experience from both primary and secondary end-users. Extend the above-mentioned guidelines by the following items to accurately reflect the context of evaluations of individually tailored DHT-enhanced training concepts (☑-item 42):

First, together with the project reference committee, work out and agree on a *traffic light system-based assessment framework* with predetermined progression criteria (☑-item 42a), as elaborating in more detail under ☑-item 15 which laid the foundation for this step. To ensure transparency in the interpretation of the results on feasibility and user experience, we recommend the traffic light system-based assessment framework be published (as part of a study protocol) and pre-registered (e.g., on the Open Science Framework) before starting data collection.

Second, in agreement with the MIDE-framework [13], systematically evaluate and report on *technology performance* (☑-item 42b; e.g., downtime, type and frequency of occurrence of technical problems, effectiveness of supportive strategies for end-users, etc.) to provide a robust basis for further iterative DHT refinements.

#### 4.3.8 Phases 7^+^: Efficacy, (Cost-)Effectiveness, and Implementation Evaluation

Finally, evaluate the efficacy, (cost-)effectiveness, and implementation of the resulting DHT-enhanced training concept, depending on the context of the project (☑-item 44). In any context, we recommend moving beyond efficacy evaluations to address the lingering evidence-to-practice gap – which is in line with the recommendations by Oluwatoyosi et al. (2020) [70]. This could be achieved by effectiveness and implementation studies following efficacy trials, or with hybrid effectiveness-implementation designs [71-73]. Moreover, assess the DHT-enhanced training concept’s value (risk-benefit profile, cost-effectiveness) for healthcare systems to provide data that allow informed decisions on the uptake of such approaches in future clinical practice guidelines.

For these evaluations, we recommend following the UK Medical Research Council guidance [7] for efficacy and effectiveness evaluations, the PRACTical planning for Implementation and Scale-up guide [34] for implementation evaluations, and the World Health Organization guide [74] for cost-effectiveness analyses (☑-item 44). Like in phase 6, expand the above-mentioned guidelines to accurately reflect the context of evaluations of individually tailored DHT-enhanced training concepts (☑-item 45) as follows.

First, while RCTs are considered the gold standard for evaluating efficacy and effectiveness [75], consider *alternative study designs* that may better accommodate the individualized and adaptive nature of tailored DHT-enhanced training [76] (☑-item 45a). Nahum-Shani et al. (2022) [77] provide a pragmatic framework for selecting such alternative designs that are particularly relevant when interventions are multicomponent or tailored and when there are research questions regarding the timing, sequencing, or responsiveness of different intervention components. For example, a factorial design is a type of randomized trial in which two or more independent variables are manipulated simultaneously, allowing researchers to assess both the main effects of each factor and potential interactions between them. This makes it an efficient approach for studying multiple intervention components within a single trial. Furthermore, the Sequential Multiple Assignment Randomized Trial design includes multiple randomizations, enabling participants to be re-randomized based on their response or adherence to the initial treatment, which is particularly useful when behaviors or conditions evolve slowly over time. In contrast, Micro-Randomized Trials involve frequent, often daily, randomizations to assess the short-term effects of brief interventions – such as prompts or notifications – on rapidly changing outcomes. Micro-Randomized Trials are valuable for real-time optimization and adaptation of DHT-enhanced training. Despite the promise of these alternative approaches in addressing the need for tailoring and adaptability in digital health, they remain underutilized in health promotion and disease prevention interventions [77].

Second, continue with *mixed methods approach* also at this stage of a project. Specifically, collect complementary participant-reported outcomes and qualitative data on perceived efficacy/effectiveness and acceptance of the implementation of primary and secondary end-users, as well as suggestions on how these could be (further) improved (☑-item 45b). These evaluations can provide insights not ascertainable through physiological, laboratory, clinician-reported, observer-reported, or performance outcomes alone. These evaluations can thereby bring additional value and optimize the impact on clinical practice and health policy. [78]

## 5. Discussion

### 5.1 Principal Results

This methodological guideline development study gradually co-created consensus-based methodological best practice recommendations for multidisciplinary expert teams coordinating projects on individually tailored DHT-enhanced training concepts. It is applicable to any type(s) of end-user(s), exercise type(s), intended context(s) of (e.g., primary healthcare, community health services, telemedicine), and overarching goal(s) (e.g., health promotion, primary through tertiary disease prevention; including rehabilitation).

### 5.2 Strengths and Limitations

This project invested considerable time and effort in collaboratively working out a consensus on the best practice recommendations – which was driven by a project team that spanned 5 research groups across 4 countries with 10 researchers representing a range of career stages, multidisciplinary backgrounds, and research interests. Nevertheless, we acknowledge that the key limitations of our approach were (i) the reliance on group decision-making and consensus-finding procedures involving (ii) a relatively small number of researchers. Given the project’s scope, we deliberately chose to develop this proposal within a group of experts with hands-on experience in applying the methodological frameworks we sought to combine and refine to create a more holistic guideline specific to individually tailored DHT-enhanced training. This approach enabled extensive in-depth critical discussion and creative brainstorming to propose, discuss, and develop a consensus on new best practice recommendations. We believe that this method is well suited to our research aims, offering richer dialog than more structured techniques such as the (modified) Delphi method [79] – which was necessary given the comprehensiveness of this work. Furthermore, the literature suggests that 10 - 15 participants may suffice when the background of such a consensus process is homogeneous [79], which applies to our research goal and methods, thereby providing justification for the number of involved experts.

Nevertheless, we recognize inherent limitations of group decision-making, including the potential influence of dominant individuals, the risk of tangential discussions, and group pressure for compromise. All of these limitations could be mitigated through alternative methods that incorporate anonymity, controlled feedback, and statistical aggregation of responses. [79] Therefore, we advocate for future iterations to collaboratively refine and consolidate this guideline through broader expert, patient, and public involvement. A (modified) Delphi method [79] – which is recommended [80, 81] but underutilized [82] in the related field of development of health research reporting guidelines – or continued co-creation procedures could be particularly valuable in this regard.

In addition to the above limitations, we acknowledge that all involved researchers are affiliated with European universities. Broader expert involvement with more diverse geographical, socio-cultural, and institutional backgrounds – particularly from low- and middle-income countries, nonacademic sectors, and underrepresented communities – would strengthen the continued refinement and consolidation of such broadly applicable best practice recommendations. Nevertheless, our consensus-finding process involved researchers with diverse educational backgrounds, including health sciences and technology, physiotherapy, movement sciences, and neuropsychology, as well as dual appointments bridging academic research with (i) clinical practice and (ii) leadership roles in technology development companies. This diversity sparked numerous critical discussions around the terminology used in different contexts, which in turn facilitated the development of a harmonized terminology that enhances the guideline’s applicability across the various interest-holders it is intended to serve.

### 5.3 Comparison with Prior Work

The *Co-Develop-IT guideline* is an extension of the MIDE - Framework [13] and its refined methodology from the ‘Brain-IT’ project [14]. It integrates the MRC guidance [7] and the Generative Co-Design Framework for Healthcare Innovation [11], aspects to inform the design of new DHTs from the NASSS framework [12] and PRACTIS - Guide [34]. Given that the *Co-Develop-IT guideline* integrated and refined these established methodological frameworks, it can be seen in part as a unified approach combining the advantages of previous frameworks that are expanded with refined recommendations specifically for the context of individually tailored DHT-enhanced training. However, the main novelty of this proposal lies in the introduction of multiple aspects to advance current best practices in the field.

*First*, the *Co-Develop-IT guideline* provides *five distinct preparatory contextual research phases preceding generative co-design and development* to establish a more robust conceptual foundation to guide targeted co-development towards successful implementation. These novel phases are dedicated to, among others, (i) harmonizing interests and clarifying the level of involvement of all contributors (particularly between research, industry, healthcare, and end-users); (ii) aligning co-design procedures with expectations and requirements of relevant interest-holders; (iii) collaboratively delineating clear strategies to monitor project progression; and (iv) facilitating implementation, scalability, and sustainability of the solutions to be developed. Regarding the latter, one of the key merits of the *Co-Develop-IT guideline* is that the implementation requirements for DHT-enhanced training are considered alongside its design requirements prior to launching and continuously during the co-development and evaluation process. Such extensive preparatory contextual research steps deviate from the currently conventional approach and extend best practice recommendations from earlier methodological frameworks. The resulting rigorous methodological approach establishes a solid foundation that facilitates the transfer of the DHT-enhanced training concept into real-world applications and clinical practice, promoting its continued use beyond project completion.

*Second*, the *Co-Develop-IT guideline* supports *harmonizing the interests of different contributors* (particularly between research, industry, healthcare, and end-users). This is done through definitions of minimum requirements for interest-holder involvement in each project step and recommendations for systematically defining their specific roles and responsibilities in the project.

*Finally*, the *Co-Develop-IT guideline* provides a corresponding *checklist with an explanation and elaboration section* to harmonize its interpretation and guide effective implementation. By briefly outlining two application examples in ongoing projects along with the guideline, we provide initial proof of concept for its practical applicability and reflect on its relevance and flexibility with the goal of strengthening the credibility and future uptake of the *Co-Develop-IT guideline* as well as the DHT-enhanced training concepts that will eventually be developed following this guideline. However, the application examples also highlighted that the *Co-Develop-IT guideline* would benefit from continued testing in different co-development projects and in different contexts to further refine and consolidate it.

### 5.4 Implications for Research

While collaborative research practices are generally considered good practices, with patient and public involvement nowadays a routine section on most grant proposals, concerns have been raised about whether the costs of collaborative research practices outweigh the benefits for health research [83]. Specifically, Oliver et al. (2019) provided an overview of the potential risks and costs of co-production and related participatory research practices such as co-design. Such risks and costs include, but are not limited to, increased time and resource demands, emotional and professional burdens on both researchers and interest holders, challenges in managing conflicting expectations, risks to academic credibility, and potential dilution of research quality due to compromises made during collaboration. To overcome these risks, they recommend *“[…] clarifying exactly which outcomes are required for whom for any particular piece of research [and] selecting strategies specifically designed to enable these outcomes to be achieved, and properly evaluated.”* [83] Furthermore, they called for a cautious approach to co-development in the absence of strong evidence supporting its process and impact [83]. While there is now moderate quality evidence that co-creation is effective by resulting in interventions that have the intended effect in the context of the secondary prevention of non-communicable diseases [84], recent research agrees that standardized definitions and reporting frameworks for co-development are needed to facilitate comparisons and ensure consistency [84, 85].

The *Co-Develop-IT guideline* provides a proposal for best practice recommendations for implementing, among others, the above-mentioned recommendations. Specifically, it entails collaboratively – with all relevant interest-holders – establishing (i) project checkpoints consisting of a traffic light system-based assessment framework with quantitative thresholds and clear progression criteria, and (ii) agreeing on the weighting of how inputs from different interest-holders should be prioritized. Furthermore, the *Co-Develop-IT checklist* guided standardized reporting of co-development projects and defines the minimum level of participatory research involvement for each item. It thereby provides guidance in which projects steps which interest-holders should be involved to what extent to ensure successful goal achievement. These strategies support projects in finding a good balance between the benefits and increased costs of collaborative research practices. Ultimately, adopting these recommendations will help advance the field to get the most out of such research projects for advancing health research that is disseminated and implemented in real-world settings.

### 5.5 Implications for Practice

Credible DHT-enhanced interventions and related DHTs should be evidence-based and grounded in health research that provides data to support whether these products achieve their intended outcomes. However, a recent review revealed a significant gap between commercial offerings and scientific validation of DHT-enhanced lifestyle interventions in the example of dementia prevention. Specifically, all reviewed studies focused on proxy measures of dementia prevention, and none demonstrated any change in the targeted condition or risk factors. [86] This observation is likely to be transferable to other use cases and highlights the need for rigorous empirical validation – rather than solely equivalence claims – to provide evidence to support the often-advertised claims on clinical effectiveness of such DHT-enhanced interventions. Following the *Co-Develop-IT guideline* in future projects will result in the (i) co-development of well-grounded DHTs and DHT-enhanced training concepts that are well aligned with all interest-holders’ requirements, (ii) provision of robust data whether the overarching goals of the specific DHT-enhanced training concept are supported by appropriate scientific evidence, and (iii) proof-of-concept for the success of implementation of the resulting DHT-enhanced training concepts. Thereby, the *Co-Develop-IT guideline* systematically guides projects towards making a real-world impact.

## 6. Conclusions

The *Co-Develop-IT guideline* proposes best practices for how participatory research and patient and public involvement may be efficiently implemented and structured to benefit the establishment of individually tailored DHT-enhanced training concepts. Given that this guideline integrated and refined previous methodological frameworks [7, 11-14, 34], it can be seen in part as a unified approach combining the advantages of previous frameworks that are expanded with refined recommendations specifically for the context of individually tailored DHT-enhanced training. However, its main novelty lies in guiding the structured establishment of a more robust conceptual foundation through extensive preparatory contextual research phases, aimed at better targeting tailored co-development efforts toward successful implementation. We advocate for the continued refinement and consolidation of this methodological guideline to help the field strike a better balance between maximizing the benefits and mitigating the increased resource demands of collaborative research practices – ultimately maximizing its real-world impact. To encourage future initiatives of this kind, we are providing a platform on the Open Science Framework [87]. We encourage the use of this platform to engage in critical discussions about experiences of using the *Co-Develop-IT guideline* to guide projects and to document efforts to test, refine, and consolidate the guideline further.

## 7. Declarations

### 7.1 Acknowledgments

The authors have nothing to declare.

### 7.4 Data Availability

All data analyzed in this study is included in the article and supplementary files. Further inquiries can be directed to the corresponding author or Research Data Office at Karolinska Institutet.

### 7.5 Author Contributions

*Conceptualization:* PM initiated, led, and coordinated the project, which began in June 2024. PM developed the initial project proposal with feedback from EF. PM and EF jointly selected and invited members to the expert panel.

#### *Methodology:* All authors contributed feedback on the project proposal and participated in its revision.

*Investigation & Data Curation:* All authors participated in hybrid meetings to collaboratively develop this consensus-based methodological guideline between August 2024 to February 2025. PM took meeting protocols, synthesized the outcomes of each meeting, and drafted the corresponding sections of the guideline, checklist, and explanation and elaboration documents. All authors provided written feedback and revisions between meetings.

#### *Visualization:* The *Co-Develop-IT guideline* figure was co-created by PM and LH.

*Writing – Original Draft:* PM drafted the main manuscript and Supplementary Files 1, 2, and 5. The first application example (Supplementary File 3) was developed by PM, EF, AW, BL, and FA. The second application example (Supplementary File 4) was developed by LH and MR.

*Writing – Review & Editing:* All authors contributed to revisions of the manuscript and supplementary files and approved the final submitted version.

## 8. Abbreviations

Co-Develop-IT: Co-design, Development, and Evaluation of Individually Tailored Technology-Enhanced Training and Rehabilitation Concepts
DHT: digital health technologies
EQUATOR: Enhancing the QUAlity and Transparency Of health Research
MIDE: Multidisciplinary Iterative Design of Exergames
MRC: Medical Research Council
NASSS: nonadoption, abandonment, scale-up, spread, and sustainability
PD: Parkinson’s disease
PRACTIS: PRACTical planning for Implementation and Scale-up
RCT: randomized controlled trial
SWOT: Strength, Weakness, Opportunities, and Threats

## References

1. Gajardo Sánchez AD, Murillo-Zamorano LR, et al. Gamification in Health Care Management: Systematic Review of the Literature and Research Agenda. SAGE Open. 2023;13(4):21582440231218834. doi: 10.1177/21582440231218834.

2. Manser P, de Bruin ED, et al. Beyond “Just” Fun: The Role of Exergames in Advancing Health Promotion and Disease Prevention. Neuroscience & Biobehavioral Reviews. 2025:106260. doi: 10.1016/j.neubiorev.2025.106260.

3. Maggio MG, Baglio F, et al. The overlooked role of exergames in cognitive-motor neurorehabilitation: a systematic review. npj Digital Medicine. 2025;8(1):419. doi: 10.1038/s41746-025-01843-4.

4. Singh B, Ahmed M, et al. A systematic umbrella review and meta-meta-analysis of eHealth and mHealth interventions for improving lifestyle behaviours. npj Digital Medicine. 2024;7(1):179. doi: 10.1038/s41746-024-01172-y.

5. World Health Organization. Recommendations on digital interventions for health system strengthening. 2019 URL: https://www.who.int/publications/i/item/9789241550505.

6. Herold F, Theobald P, et al. The Best of Two Worlds to Promote Healthy Cognitive Aging: Definition and Classification Approach of Hybrid Physical Training Interventions. JMIR Aging. 2024;7:e56433. doi: 10.2196/56433.

7. Skivington K, Matthews L, et al. A new framework for developing and evaluating complex interventions: update of Medical Research Council guidance. BMJ. 2021;374:n2061. doi: 10.1136/bmj.n2061.

8. Booth V, Hood-Moore V, et al. Systematic scoping review of frameworks used to develop rehabilitation interventions for older adults. BMJ Open. 2019;9(2):e024185. doi: 10.1136/bmjopen-2018-024185.

9. Moore G, Wilding H, et al. Participatory Methods to Engage Health Service Users in the Development of Electronic Health Resources: Systematic Review. J Participat Med. 2019;11(1):e11474. doi: 10.2196/11474.

10. Abras C, Maloney-Krichmar D, Preece J. User-centered design. Bainbridge, W Encyclopedia of Human-Computer Interaction Thousand Oaks: Sage Publications, vol 4. 2004 URL: https://aim.johnkeston.com/wp-content/uploads/2012/01/User-centered_design_encyclopedia_chapter.pdf.

11. Bird M, McGillion M, et al. A generative co-design framework for healthcare innovation: development and application of an end-user engagement framework. Research Involvement and Engagement. 2021;7(1):12. doi: 10.1186/s40900-021-00252-7.

12. Greenhalgh T, Wherton J, et al. Beyond Adoption: A New Framework for Theorizing and Evaluating Nonadoption, Abandonment, and Challenges to the Scale-Up, Spread, and Sustainability of Health and Care Technologies. Journal of Medical Internet Research. 2017;19(11):e367. doi: 10.2196/jmir.8775.

13. Li Y, Muñoz J, et al. Multidisciplinary Iterative Design of Exergames (MIDE): A Framework for Supporting the Design, Development, and Evaluation of Exergames for Health. International Conference on Human-Computer Interaction: Springer; 2020. p. 128–147.doi: 10.1007/978-3-030-50164-8_9

14. Manser P, de Bruin ED. Making the Best Out of IT: Design and Development of Exergames for Older Adults With Mild Neurocognitive Disorder - A Methodological Paper. Front Aging Neurosci. 2021;13:734012. doi: 10.3389/fnagi.2021.734012.

15. Manser P, de Bruin ED. “Brain-IT”: Exergame training with biofeedback breathing in neurocognitive disorders. Alzheimer’s & dementia. 2024;20(7):4747–4764. doi: 10.1002/alz.13913.

16. Huber SK, Manser P, de Bruin ED. PEMOCS: theory derivation of a concept for PErsonalized MOtor-Cognitive exergame training in chronic Stroke—a methodological paper with an application example. Frontiers in Sports and Active Living. 2024;6. doi: 10.3389/fspor.2024.1397949.

17. Herold F, Müller P, et al. Dose–Response Matters! – A Perspective on the Exercise Prescription in Exercise– Cognition Research. Frontiers in psychology. 2019;10(2338). doi: 10.3389/fpsyg.2019.02338.

18. Vaughn LM, Jacquez F. Participatory research methods–choice points in the research process. Journal of participatory research methods. 2020;1(1). doi: 10.35844/001c.13244.

19. Network E: Enhancing the QUAlity and Transparency Of health Research. https://www.equator-network.org/ (2025). Accessed May 30 2025.

20. Boers M, Rochereau A, et al. Classification grid and evidence matrix for evaluating digital medical devices under the European union landscape. npj Digital Medicine. 2025;8(1):304. doi: 10.1038/s41746-025-01697-w.

21. Manser P, Poikonen H, de Bruin ED. Feasibility, usability, and acceptance of “Brain-IT”—A newly developed exergame-based training concept for the secondary prevention of mild neurocognitive disorder: a pilot randomized controlled trial. Frontiers in Aging Neuroscience. 2023;15. doi: 10.3389/fnagi.2023.1163388.

22. Wallin A, Franzén E, et al. A highly challenging balance training intervention for people with multiple sclerosis: a feasibility trial. Pilot and Feasibility Studies. 2023;9(1):41. doi: 10.1186/s40814-023-01265-7.

23. Cairns P, Pinker I, et al. Empathy maps in communication skills training. The Clinical Teacher. 2021;18(2):142–146. doi: 10.1111/tct.13270.

24. Rohner SL, Stadtmann MP, et al. Co-creation for the development and implementation of a competence centre for mental health in Eastern Switzerland: a participatory approach. BMC Psychiatry. 2025;25(1):254. doi: 10.1186/s12888-025-06703-9.

25. Mental Health Europe. Guidelines for Co-Creation in Mental Health. 2023 URL: https://www.mentalhealtheurope.org/wp-content/uploads/2024/04/Mental-Health-Europes-Guidelines-for-Cocreation.pdf.

26. Izquierdo M, de Souto Barreto P, et al. Global consensus on optimal exercise recommendations for enhancing healthy longevity in older adults (ICFSR). The Journal of nutrition, health and aging. 2025:100401. doi: 10.1016/j.jnha.2024.100401.

27. American College of Sports Medicine. ACSM’s Guidelines for Exercise Testing and Prescription. Wolters Kluwer; 2025 URL: https://acsm.org/education-resources/books/guidelines-exercise-testing-prescription/.

28. Kleim Jeffrey A, Jones Theresa A. Principles of Experience-Dependent Neural Plasticity: Implications for Rehabilitation After Brain Damage. Journal of Speech, Language, and Hearing Research. 2008;51(1):S225–S239. doi: 10.1044/1092-4388(2008/018).

29. Kleim JA. Neural plasticity and neurorehabilitation: Teaching the new brain old tricks. Journal of Communication Disorders. 2011;44(5):521–528. doi: 10.1016/j.jcomdis.2011.04.006.

30. Maier M, Ballester BR, Verschure PFMJ. Principles of Neurorehabilitation After Stroke Based on Motor Learning and Brain Plasticity Mechanisms. Frontiers in Systems Neuroscience. 2019;13. doi: 10.3389/fnsys.2019.00074.

31. Rhodes RE, McEwan D, Rebar AL. Theories of physical activity behaviour change: A history and synthesis of approaches. Psychology of Sport and Exercise. 2019;42:100–109. doi: 10.1016/j.psychsport.2018.11.010.

32. Herold F, Hamacher D, et al. Thinking While Moving or Moving While Thinking - Concepts of Motor-Cognitive Training for Cognitive Performance Enhancement. Frontiers in Aging Neuroscience. 2018;10(228). doi: 10.3389/fnagi.2018.00228.

33. Raichlen DA, Alexander GE. Adaptive Capacity: An Evolutionary Neuroscience Model Linking Exercise, Cognition, and Brain Health. Trends in Neurosciences. 2017;40(7):408–421. doi: 10.1016/j.tins.2017.05.001.

34. Koorts H, Eakin E, et al. Implementation and scale up of population physical activity interventions for clinical and community settings: the PRACTIS guide. International Journal of Behavioral Nutrition and Physical Activity. 2018;15(1):51. doi: 10.1186/s12966-018-0678-0.

35. Cheval B, Boisgontier MP. The Theory of Effort Minimization in Physical Activity. Exercise and Sport Sciences Reviews. 2021;49(3). doi: 10.1249/JES.0000000000000252.

36. Gerber M, Cheval B, et al. Psycho-physiological foundations of human physical activity behavior and motivation: Theories, systems, mechanisms, evolution, and genetics. Physiological Reviews. 2025. doi: 10.1152/physrev.00021.2024.

37. Wulf G, Lewthwaite R. Optimizing performance through intrinsic motivation and attention for learning: The OPTIMAL theory of motor learning. Psychon Bull Rev. 2016;23(5):1382–1414. doi: 10.3758/s13423-015-0999-9.

38. El Kirat H, van Belle S, et al. Behavioral change interventions, theories, and techniques to reduce physical inactivity and sedentary behavior in the general population: a scoping review. BMC Public Health. 2024;24(1):2099. doi: 10.1186/s12889-024-19600-9.

39. Hamasaki T, Briand C, et al. Digital health technology adoption factors: a rapid review of systematic reviews and checklist development. Disability and Rehabilitation: Assistive Technology. 2025:1–18. doi: 10.1080/17483107.2025.2526175.

40. Burns PB, Rohrich RJ, Chung KC. The Levels of Evidence and Their Role in Evidence-Based Medicine. Plastic and Reconstructive Surgery. 2011;128(1). doi: 10.1097/PRS.0b013e318219c171.

41. Gavi B, Daniel H. Analysing Health Communication - Discourse Approaches. Springer Nature Link: Springer Nature; 2021 URL: https://link.springer.com/book/10.1007/978-3-030-68184-5.

42. Marta B, Maria Laura I, Jaume R. Healthcare in the Digital Age - Perspectives for Sustainable Innovation and Assessment. Springer Nature Link: Springer Nature; 2025 URL: 10.1007/978-981-96-1437-0.

43. Torre MM, Temprado J-J. Effects of Exergames on Brain and Cognition in Older Adults: A Review Based on a New Categorization of Combined Training Intervention. Frontiers in Aging Neuroscience. 2022;14. doi: 10.3389/fnagi.2022.859715.

44. Farr M. Power dynamics and collaborative mechanisms in co-production and co-design processes. Critical Social Policy. 2017;38(4):623–644. doi: 10.1177/0261018317747444.

45. Wilson C. Chapter 2 - Brainwriting. Brainstorming and Beyond. Boston: Morgan Kaufmann; 2013 URL: https://www.sciencedirect.com/science/article/pii/B9780124071575000026.

46. Gray D, Brown S, Macanufo J. Gamestorming: A playbook for innovators, rulebreakers, and changemakers. " O’Reilly Media, Inc."; 2010 URL: https://gamestorming.com/.

47. Chang W-L, Shao Y-C. Co-creating User Journey Map – A Systematic Approach to Exploring Users’ Day-to-Day Experience in Participatory Design Workshops. In: Kurosu M, Hashizume A, editors. Human-Computer Interaction. Cham: Springer Nature Switzerland; 2023. p. 3–17.doi: 10.1007/978-3-031-35596-7_1

48. Segura E, Vidal L, Rostami A. Bodystorming for movement-based interaction design. Human Technology. 2016;12(2):193–251. doi: 10.17011/ht/urn.201611174655.

49. Schleicher D, Jones P, Kachur O. Bodystorming as embodied designing. Interactions. 2010;17(6):47–51. doi: 10.1145/1865245.1865256.

50. Mahatody T, Sagar M, Kolski C. State of the Art on the Cognitive Walkthrough Method, Its Variants and Evolutions. International Journal of Human–Computer Interaction. 2010;26(8):741–785. doi: 10.1080/10447311003781409.

51. Mayring P. Qualitative content analysis: A step-by-step guide. 2021 URL: https://uk.sagepub.com/en-gb/eur/qualitative-content-analysis/book269922.

52. Krippendorff K. Content Analysis: An Introduction to Its Methodology. Fourth Edition ed. Thousand Oaks, California: 2019 https://methods.sagepub.com/book/content-analysis-4e.

53. Mayring P. Qualitative content analysis: theoretical foundation, basic procedures and software solution. 2014. doi: 10.1007/978-94-017-9181-6_13.

54. Peters S, Guccione L, et al. Evaluation of research co-design in health: a systematic overview of reviews and development of a framework. Implementation Science. 2024;19(1):63. doi: 10.1186/s13012-024-01394-4.

55. Schrepp M, Thomaschewski J. Design and validation of a framework for the creation of user experience questionnaires. IJIMAI. 2019;5(7):88–95. doi: 10.9781/ijimai.2019.06.006.

56. Winter D, Hinderks A, et al. Welche UX Faktoren sind für mein Produkt wichtig? Mensch und Computer 2017-Usability Professionals: Gesellschaft für Informatik eV; 2017.doi: 10.18420/muc2017-up-0002

57. Winter D, Schrepp M, Thomaschewski J. Faktoren der User Experience-Systematische Übersicht über produktrelevante UX-Qualitätsaspekte. Mensch und Computer 2015–Usability Professionals: De Gruyter Oldenbourg; 2015. p. 33–41.doi: 10.1515/9783110443882-005

58. Perez FMP, Bellei EA, et al. Decoding user experience in exergames: A systematic scoping review of assessment methods. MethodsX. 2025;14:103054. doi: 10.1016/j.mex.2024.103054.

59. Johnson RM. User Experience Research and Usability of Health Information Technology. Technical Communication Quarterly. 2025:1–4. doi: 10.1080/10572252.2025.2455553.

60. Campbell JL. User Experience Research and Usability of Health Information Technology. Auerbach Publications; 2024 URL: 10.1201/9781003460886.

61. Guggenberger B, Jocham AJ, et al. Instrumental Validity of the Motion Detection Accuracy of a Smartphone-Based Training Game. International Journal of Environmental Research and Public Health. 2021;18(16). doi: 10.3390/ijerph18168410.

62. Kaiser W, de Bruin ED, Manser P. Domain-Specific Evaluation of Exergame Metrics Among Older Adults With Mild Neurocognitive Disorder: Secondary Analysis of 2 Randomized Controlled Trials. JMIR Serious Games. 2025;13:e65878. doi: 10.2196/65878.

63. Guimarães V, Sousa I, et al. Using shoe-mounted inertial sensors and stepping exergames to assess the motor-cognitive status of older adults: A correlational study. DIGITAL HEALTH. 2023;9:20552076231167001. doi: 10.1177/20552076231167001.

64. Konstantinidis EI, Bamidis PD, et al. Physical Training In-Game Metrics for Cognitive Assessment: Evidence from Extended Trials with the Fitforall Exergaming Platform. Sensors. 2021;21(17). doi: 10.3390/s21175756.

65. Petsani D, Konstantinidis E, et al. Digital Biomarkers for Well-being Through Exergame Interactions: Exploratory Study. JMIR Serious Games. 2022;10(3):e34768. doi: 10.2196/34768.

66. Litz E, Ball C, et al. Validation of a Motor-Cognitive Assessment for a Stepping Exergame in Older Adults: Use of Game-Specific, Internal Data Stream. Games Health J. 2020;9(2):95–107. doi: 10.1089/g4h.2019.0081.

67. Wiloth S, Lemke N, et al. Validation of a Computerized, Game-based Assessment Strategy to Measure Training Effects on Motor-Cognitive Functions in People With Dementia. JMIR Serious Games. 2016;4(2):e12. doi: 10.2196/games.5696.

68. Eldridge SM, Lancaster GA, et al. Defining Feasibility and Pilot Studies in Preparation for Randomised Controlled Trials: Development of a Conceptual Framework. PLoS One. 2016;11(3):e0150205. doi: 10.1371/journal.pone.0150205.

69. O’Cathain A, Hoddinott P, et al. Maximising the impact of qualitative research in feasibility studies for randomised controlled trials: guidance for researchers. Pilot and Feasibility Studies. 2015;1(1):32. doi: 10.1186/s40814-015-0026-y.

70. Oluwatoyosi BAO, Rachel SR, Ross CB. Dissemination and implementation research in sports and exercise medicine and sports physical therapy: translating evidence to practice and policy. BMJ Open Sport & Exercise Medicine. 2020;6(1):e000974. doi: 10.1136/bmjsem-2020-000974.

71. Landes SJ, McBain SA, Curran GM. An introduction to effectiveness-implementation hybrid designs. Psychiatry Res. 2019;280:112513. doi: 10.1016/j.psychres.2019.112513.

72. Curran GM, Bauer M, et al. Effectiveness-implementation Hybrid Designs: Combining Elements of Clinical Effectiveness and Implementation Research to Enhance Public Health Impact. Medical Care. 2012;50(3). doi: 10.1097/MLR.0b013e3182408812.

73. Curran GM, Landes SJ, et al. Reflections on 10 years of effectiveness-implementation hybrid studies. Frontiers in Health Services. 2022;Volume 2 - 2022. doi: 10.3389/frhs.2022.1053496.

74. Edejer TT-T, Edejer TT-T. Making choices in health: WHO guide to cost-effectiveness analysis. World Health Organization; 2003 URL: https://iris.who.int/handle/10665/42699.

75. Hariton E, Locascio JJ. Randomised controlled trials – the gold standard for effectiveness research. BJOG: An International Journal of Obstetrics & Gynaecology. 2018;125(13):1716–1716. doi: 10.1111/1471-0528.15199.

76. Daniels K, Quadflieg K, Bonnechère B. Mobile health interventions for active aging: a systematic review and meta-analysis on the effectiveness of physical activity promotion. mHealth. 2025;11. doi: 10.21037/mhealth-24-41.

77. Nahum-Shani I, Dziak JJ, Wetter DW. MCMTC: A Pragmatic Framework for Selecting an Experimental Design to Inform the Development of Digital Interventions. Frontiers in Digital Health. 2022;Volume 4 - 2022. doi: 10.3389/fdgth.2022.798025.

78. Crossnohere NL, Schuster ALR, et al. Patient-reported outcome measures add value as clinical trial endpoints. Nature Medicine. 2025. doi: 10.1038/s41591-025-03906-1.

79. Taylor E. We Agree, Don’t We? The Delphi Method for Health Environments Research. HERD: Health Environments Research & Design Journal. 2020;13(1):11–23. doi: 10.1177/1937586719887709.

80. Moher D, Schulz KF, et al. Guidance for Developers of Health Research Reporting Guidelines. Plos Medicine. 2010;7(2):e1000217. doi: 10.1371/journal.pmed.1000217.

81. Black N, Murphy M, et al. Consensus Development Methods: A Review of Best Practice in Creating Clinical Guidelines. Journal of Health Services Research & Policy. 1999;4(4):236–248. doi: 10.1177/135581969900400410.

82. Banno M, Tsujimoto Y, Kataoka Y. The majority of reporting guidelines are not developed with the Delphi method: a systematic review of reporting guidelines. Journal of Clinical Epidemiology. 2020;124:50–57. doi: 10.1016/j.jclinepi.2020.04.010.

83. Oliver K, Kothari A, Mays N. The dark side of coproduction: do the costs outweigh the benefits for health research? Health Research Policy and Systems. 2019;17(1):33. doi: 10.1186/s12961-019-0432-3.

84. Anieto EM, Dall PM, et al. The effectiveness of co-created lifestyle interventions in improving health behaviour, physical and mental health in adults with non-communicable diseases: A systematic review with meta-analysis. Public health. 2025;248:105929. doi: 10.1016/j.puhe.2025.105929.

85. Longworth GR, de Boer J, et al. Navigating process evaluation in co-creation: a Health CASCADE scoping review of used frameworks and assessed components. BMJ Glob Health. 2024;9(7). doi: 10.1136/bmjgh-2023-014483.

86. Vinay R, Probst J, et al. Top-funded digital health companies offering lifestyle interventions for dementia prevention: Company overview and evidence analysis. PLoS One. 2025;20(5):e0323390. doi: 10.1371/journal.pone.0323390.

87. Manser P, Hardeman LES, et al. Platform for Exchanges, Refinements, and Consolidation of Co-Develop-IT: Co-design, Development, and Evaluation of Serious Individually Tailored Technology-Enhanced Training Approaches. 2025 URL: https://osf.io/w2jd3/.

